# Evolution of COVID-19 symptoms during the first 9 months after illness onset

**DOI:** 10.1101/2021.05.05.21256710

**Authors:** Elke Wynberg, Hugo van Willigen, Maartje Dijkstra, Anders Boyd, Neeltje A. Kootstra, Joost G. van den Aardweg, Marit J. van Gils, Amy Matser, Marije R. de Wit, Tjalling Leenstra, Godelieve de Bree, Menno D. de Jong, Maria Prins, the RECoVERED Study Group

## Abstract

**Background:** Few longitudinal data on COVID-19 symptoms across the full spectrum of disease severity are available. We evaluated symptom onset, severity and recovery up to nine months after illness onset.

**Methods:** The RECoVERED Study is a prospective cohort study based in Amsterdam, the Netherlands. Participants aged>18 years were recruited following SARS-CoV-2 diagnosis via the local Public Health Service and from hospitals. Standardised symptom questionnaires were completed at recruitment, at one week and month after recruitment, and monthly thereafter. Clinical severity was defined according to WHO criteria. Kaplan-Meier methods were used to compare time from illness onset to symptom recovery, by clinical severity. We examined determinants of time to recovery using multivariable Cox proportional hazards models.

**Results:** Between 11 May 2020 and 31 January 2021, 301 COVID-19 patients (167[55%] male) were recruited, of whom 99/301(32.9%) had mild, 140/301(46.5%) moderate, 30/301(10.0%) severe and 32/301(10.6%) critical disease. The proportion of symptomatic participants who reported at least one persistent symptom at 12 weeks after illness onset was greater in those with severe/critical disease (81.7%[95%CI=68.7-89.7%]) compared to those with mild or moderate disease (33.0%[95%CI=23.0-43.3%] and 63.8%[95%CI=54.8-71.5%]). Even at nine months after illness onset, almost half of all participants (42.1%[95%CI=35.6-48.5]) overall continued to report ≥1 symptom. Recovery was slower in participants with BMI≥30kg/m^2^ (HR 0.51[95%CI=0.30-0.87]) compared to those with BMI<25kg/m^2^, after adjusting for age, sex and number of comorbidities.

**Conclusions:** COVID-19 symptoms persisted for nine months after illness onset, even in those with mild disease. Obesity was the most important determinant of speed of recovery from symptoms.

## Introduction

The clinical spectrum of coronavirus disease 2019 (COVID-19), caused by severe acute respiratory syndrome coronavirus 2 (SARS-CoV-2), ranges from asymptomatic presentation to fatal illness. Although the acute symptomatology of hospitalised patients has been well documented[1-4], less is known about the symptomatology of milder cases. In addition, robust longitudinal data on the long-term persistence of symptoms and factors that may affect recovery are scarce.

A growing body of evidence suggests that post-COVID syndrome (i.e. long COVID or Post-Acute Sequelae of SARS-CoV-2 infection [PASC]) may negatively affect both individual quality of life and the economic productivity of society [5-7]. A recent cross-sectional study from Wuhan, China, for example, reported that the vast majority (76%) of previously hospitalised patients still suffered from at least one symptom, most commonly fatigue, six months after symptom onset[7].

The RECoVERED study is a prospective cohort study of individuals with SARS-CoV-2 infection across the full spectrum of disease severity. We evaluated the incidence, severity and duration of symptoms in participants with mild, moderate, severe and critical COVID-19, and examined determinants of time to recovery from symptoms.

## Methods

### Study design and participants

The RECoVERED study is an ongoing cohort study of individuals with laboratory-confirmed SARS-CoV-2 infection in Amsterdam, the Netherlands. The study aims to describe the immunological, clinical and psychosocial sequelae of SARS-CoV-2 infection. Recruitment of study participants began on 11 May 2020. Non-hospitalised participants were identified from notification data at the Public Health Service of Amsterdam (PHSA) and approached by telephone up to 7 days after SARS-CoV-2 diagnosis. Prospectively-enrolled hospitalised participants were approached on the COVID-19 wards of two hospitals in Amsterdam. A limited number of hospitalised patients were enrolled retrospectively until 30 June 2020 up to 3 months following SARS-CoV-2 diagnosis. For the present analyses we included all participants with a follow-up of at least one month by 28 February 2021.

Eligibility criteria included laboratory confirmation of SARS-CoV-2 infection by PCR, antigen testing or serology; aged 16-85 years and adequate understanding of Dutch or English. Patients residing in a nursing home prior to SARS-CoV-2 infection and those with mental disorders that would interfere with adherence to study procedures were excluded.

The RECoVERED study was approved by the medical ethical review board of the Amsterdam University Medical Centre (NL73759.018.20). All participants provided written informed consent.

### Study procedures

At enrolment and at day 7 and month 1 of follow-up, a standardised questionnaire (based on the World Health Organisation Case Report Form [8]) on the presence, start and stop dates, and severity of 18 symptoms was completed by interviewing the participant (Figure S1). Participants subsequently completed monthly questionnaires on the presence of symptoms.

Biological specimens were collected at each monthly study visit (Figure S1). Heart rate, respiratory rate (RR) and oxygen saturation (SpO_2_) were measured at enrolment and at day 7 of follow-up, or retrieved from hospital records for retrospectively-enrolled participants. Past medical history and socio-demographic data were collected during the first month of follow-up, and data on COVID-19-related complications, treatment, and investigations were collected at month 1 of follow-up and three-monthly from month 3. Self-reported data were verified with hospital electronic medical records when available.

### Definitions

Illness onset was the first day on which symptoms were experienced; for asymptomatic patients, this was defined as the date of SARS-CoV-2 diagnosis. Complete recovery was defined as resolution of all COVID-19 symptoms. As per National Institute for Health and Care Excellence (NICE) guidelines[9], the acute phase of disease was defined as the first 4 weeks after illness onset, and post-COVID syndrome as symptoms persisting at least 12 weeks after illness onset.

Clinical severity groups were defined based on WHO COVID-19 disease severity criteria[10]. Mild disease was defined as having a RR<20/min and SpO_2_>94% on room air at both enrolment and day 7 study visits; moderate disease as having a RR20-30/min, SpO_2_ 90-94% and/or receiving oxygen therapy at enrolment or day 7 study visits; severe disease as having a RR>30/min or SpO_2_<90% at enrolment or day 7 study visits; critical disease as requiring ICU admission as a result of COVID-19 at any point.

Symptom severity was measured on a four-point scale, with the exception of dyspnoea, measured using the 6-point modified Medical Research Council (mMRC) breathlessness scale[11]. Comorbidities at illness onset were those listed by the WHO as associated with severe COVID-19[12]: cardiovascular disease (CVD), diabetes mellitus (DM), chronic lung disease (CLD), severe liver disease, chronic kidney disease, immunodeficiency, cancer and/or cerebrovascular disease. Obesity was excluded from the comorbidity variable because body mass index (BMI) was defined separately, categorised in kg/m^2^ as: <25, underweight or normal weight; 25-30, overweight; >30, obese. Ethnicity was based on the participant and their parents’ country of birth [13].

Loss to follow-up (LTFU) was defined as active withdrawal from the study or two consecutive no-show appointments despite 3 attempts to establish contact. Date of LTFU was defined as the date of last contact with the participant.

### Statistical analyses

Socio-demographic and clinical characteristics of participants were compared between clinical severity groups.

The incidence proportions for 18 different symptoms at illness onset and one and four weeks after illness onset were calculated as the number of participants reporting each symptom over the total number of participants in follow-up at that point, and was compared by clinical severity group. Asymptomatic participants contributed to the denominator. Given the potential recall bias in reporting symptom onset, we restricted this analysis to prospectively-enrolled participants. For symptoms reported by >20% of participants at 12 weeks in any severity group (the most persistent symptoms), changes in self-reported symptom severity over time during the acute phase were visualised using alluvial plots, both overall and by clinical severity group.

The proportion of participants with ongoing symptoms (firstly, with at least one symptom and secondly, for each symptom separately) were estimated using Kaplan-Meier survival curves and compared by severity group using the log-rank test. The at-risk period began at illness onset and continued until symptom recovery, loss to follow up, 9 months after illness onset, or last study visit prior to 28 February 2021 (i.e. administrative censor date), whichever occurred first. Asymptomatic participants were excluded from all symptom survival analysis.

We assessed the effect of age, sex, BMI, and number of comorbidities at illness onset on time to complete recovery from symptoms using univariable and multivariable Cox proportional hazards models in which these four variables were included. Interactions between BMI and the other variables were assessed. Clinical severity was not included in the multivariable model due to collinearity with the other variables. Two sensitivity analyses were performed to explore the impact of recall bias: firstly, restricting the analysis to prospectively-enrolled participants and secondly, left-truncating participants at the date of study entry (ignoring any recovery event occurring prior to enrolment).

A p<0.05 was considered statistically significant. Statistical analyses were performed using Stata (StataCorp LLC, v.15.1) and R (RStudio, v.1.2.5033).

## Results

### Study population

Between 11 May 2020 and 31 January 2021, 301 participants were enrolled, of whom the majority (210/301;70.0%) prospectively. Of these 301 participants, 99 (33%) experienced mild, 140 (46%) moderate, 30 (10%) severe and 32 (11%) critical disease (Table 1). Participants with severe or critical disease were older than those with mild or moderate disease (p<0.001) and had higher BMI (p=0.006). CVD, CLD, DM and immunosuppression were significantly more common among participants with severe or critical COVID-19 compared to those with mild or moderate disease (Table 1). Median time from illness onset to recruitment was 8 days (IQR=5-12) for prospectively enrolled and 85 days (IQR=73-94) for retrospectively enrolled participants. Until 28 February 2021, 42 participants were lost to follow-up. One death, due to COVID-19, occurred during follow-up.

**Table 1.**
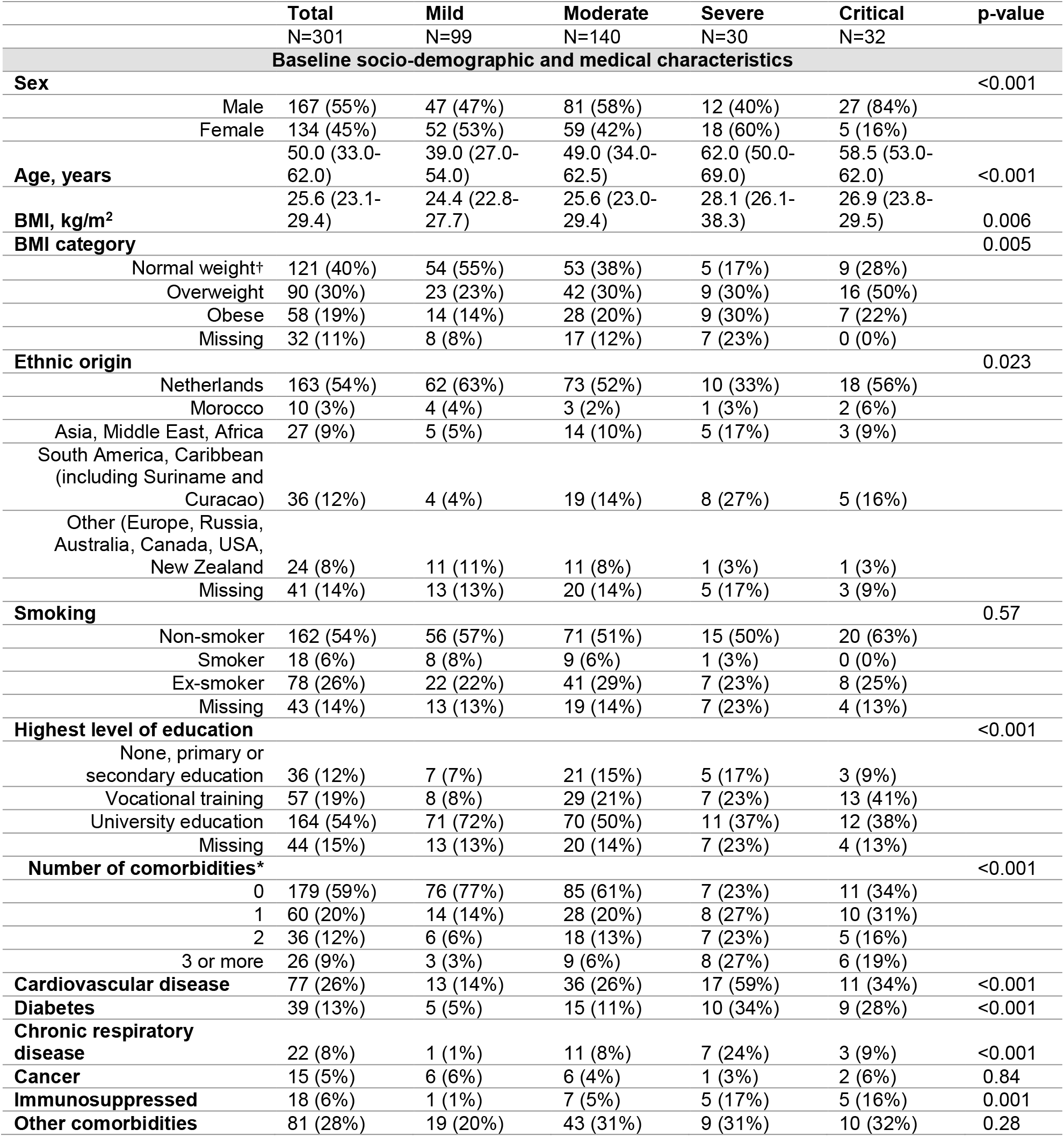
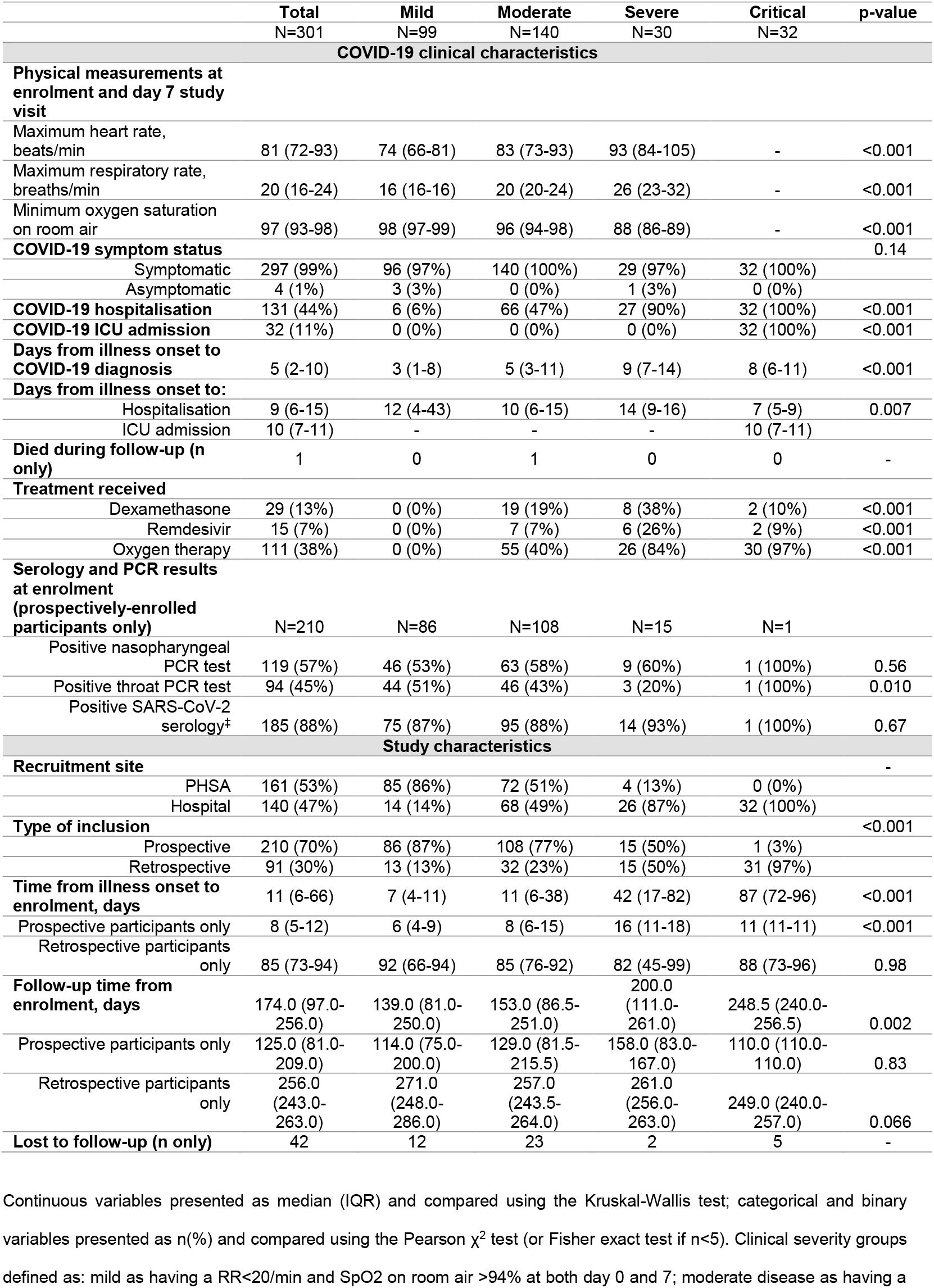
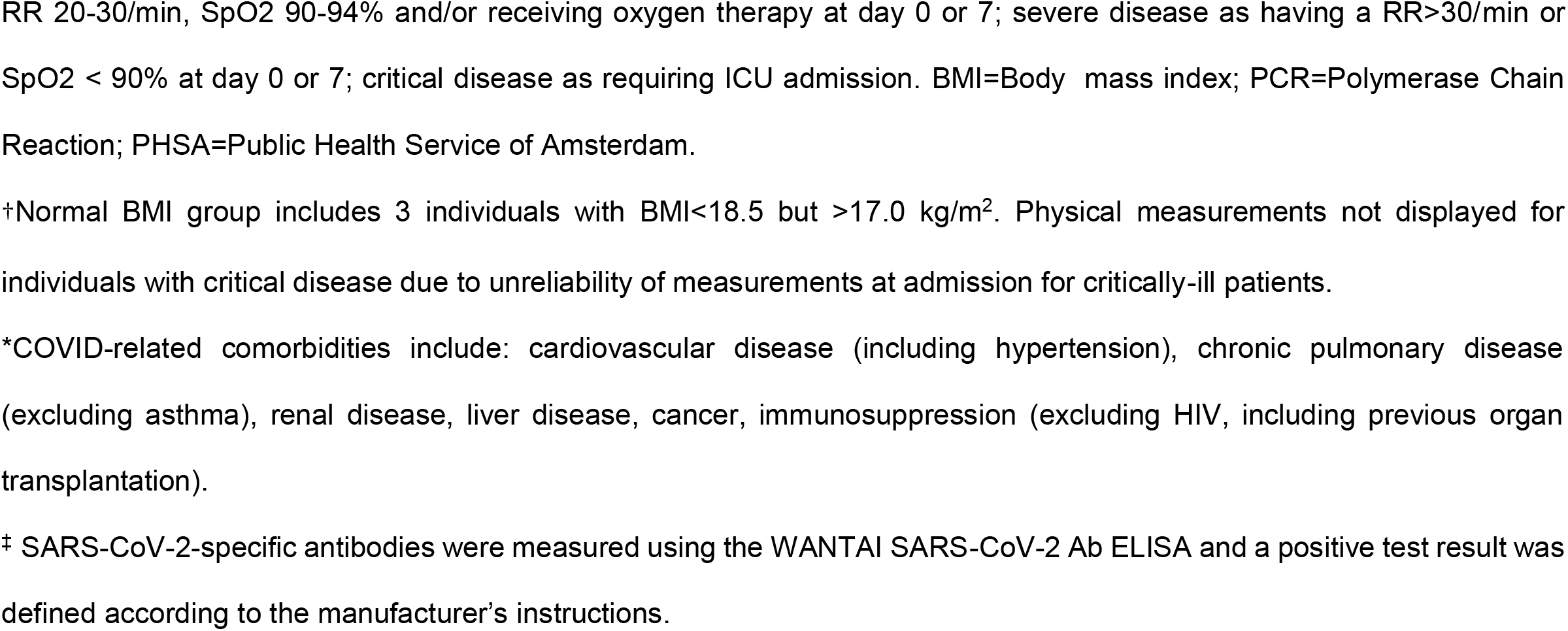
**Socio-demographic and clinical characteristics of participants of the RECoVERED study, May 2020-February 2021 in Amsterdam, the Netherlands, by clinical severity group**

### Incidence proportions and severity of symptoms during the acute phase of infection

During the acute phase of infection, fatigue and cough were the most frequently reported symptoms (Table 2). The incidence proportion for fatigue and cough as well as for fever, myalgia, loss of appetite, diarrhoea, and nausea (part of the WHO COVID-19 case definition[14]) did not differ between clinical severity groups during the acute phase (Table 2). By four weeks after illness onset, the incidence proportion for dyspnoea was higher in participants with moderate and severe/critical disease (65% and 67%, respectively; severe and critical participants combined due to small numbers) compared to participants with mild disease (43%). Within the first week after illness onset, a significantly greater proportion of patients with mild and moderate COVID-19 reported loss of smell and/or taste, headache, rhinorrhoea and sore throat compared to the severe/critical COVID-19 group and, with the exception of sore throat, this difference remained at one month after illness onset.

**Table 2.** **Incidence proportion of 18 symptoms at illness onset and within one and four weeks after illness onset (the acute phase of disease) among prospectively-recruited participants, by clinical severity group**

|  | On day of illness onset |  |  |  |  | Within one week from illness onset |  |  |  | Within four weeks from illness onset |  |  |  |
| --- | --- | --- | --- | --- | --- | --- | --- | --- | --- | --- | --- | --- | --- |
|  | All patients | Mild disease | Moderate disease | Severe/<br>Critical disease | p-value | Mild disease | Moderate disease | Severe/<br>Critical disease | p-value | Mild disease | Moderate disease | Severe/<br>Critical disease | p-value |
|  | N=210 | N=86 | N=108 | N=16 |  | N=86 | N=108 | N=16 |  | N=83 | N=105 | N=15 |  |
| **Fatigue** | 107 (51%) | 46 (53%) | 55 (51%) | 6 (38%) | 0.50 | 70 (81%) | 76 (70%) | 10 (63%) | 0.12 | 72 (87%) | 87 (83%) | 11 (73%) | 0.37 |
| **Cough** | 75 (36%) | 30 (35%) | 40 (37%) | 5 (31%) | 0.88 | 51 (59%) | 65 (60%) | 9 (56%) | 0.95 | 55 (66%) | 75 (71%) | 10 (67%) | 0.68 |
| **Loss of smell and/or taste** | 41 (20%) | 19 (22%) | 20 (19%) | 2 (13%) | 0.66 | 60 (70%) | 51 (47%) | 7 (44%) | 0.004 | 66 (80%) | 62 (59%) | 7 (47%) | 0.004 |
| **Headache** | 76 (36%) | 39 (45%) | 33 (31%) | 4 (25%) | 0.066 | 56 (65%) | 52 (48%) | 6 (38%) | 0.022 | 60 (72%) | 61 (58%) | 6 (40%) | 0.025 |
| **Rhinorrhoea** | 55 (26%) | 26 (30%) | 28 (26%) | 1 (6%) | 0.13 | 55 (64%) | 53 (49%) | 3 (19%) | 0.002 | 61 (73%) | 60 (57%) | 6 (40%) | 0.014 |
| **Myalgia** | 73 (35%) | 34 (40%) | 35 (32%) | 4 (25%) | 0.46 | 49 (57%) | 56 (52%) | 8 (50%) | 0.77 | 56 (67%) | 61 (58%) | 9 (60%) | 0.47 |
| **Fever** | 77 (37%) | 29 (34%) | 40 (37%) | 8 (50%) | 0.46 | 41 (48%) | 56 (52%) | 11 (69%) | 0.30 | 45 (54%) | 64 (61%) | 12 (75%) | 0.21 |
| **Loss of appetite** | 56 (27%) | 22 (26%) | 31 (29%) | 3 (19%) | 0.72 | 41 (48%) | 57 (53%) | 9 (56%) | 0.74 | 45 (54%) | 66 (63%) | 10 (67%) | 0.44 |
| **Dyspnoea** | 37 (18%) | 10 (12%) | 26 (24%) | 1 (6%) | 0.042 | 32 (37%) | 50 (46%) | 5 (31%) | 0.31 | 36 (43%) | 68 (65%) | 10 (67%) | 0.010 |
| **Sore throat** | 52 (25%) | 27 (31%) | 24 (22%) | 1 (6%) | 0.069 | 48 (56%) | 43 (40%) | 3 (19%) | 0.007 | 50 (60%) | 51 (49%) | 5 (33%) | 0.086 |
| **Diarrhoea** | 23 (11%) | 9 (10%) | 12 (11%) | 2 (13%) | 0.94 | 24 (28%) | 26 (24%) | 4 (25%) | 0.86 | 28 (34%) | 40 (38%) | 4 (27%) | 0.61 |
| **Chest pain** | 19 (9%) | 8 (9%) | 8 (7%) | 3 (19%) | 0.27 | 26 (30%) | 24 (22%) | 4 (25%) | 0.44 | 28 (34%) | 37 (35%) | 6 (40%) | 0.90 |
| **Nausea** | 25 (12%) | 10 (12%) | 14 (13%) | 1 (6%) | 0.85 | 20 (23%) | 32 (30%) | 3 (19%) | 0.53 | 23 (28%) | 39 (37%) | 3 (20%) | 0.23 |
| **Abdominal pain** | 17 (8%) | 8 (9%) | 9 (8%) | 0 (0%) | 0.64 | 20 (23%) | 22 (20%) | 3 (19%) | 0.90 | 26 (31%) | 30 (29%) | 2 (13%) | 0.37 |
| **Arthralgia** | 26 (12%) | 11 (13%) | 14 (13%) | 1 (6%) | 0.90 | 21 (24%) | 16 (15%) | 2 (13%) | 0.22 | 23 (28%) | 28 (27%) | 4 (27%) | 1.00 |
| **Confusion** | 15 (7%) | 10 (12%) | 5 (5%) | 0 (0%) | 0.11 | 14 (16%) | 12 (11%) | 2 (13%) | 0.54 | 16 (19%) | 21 (20%) | 3 (20%) | 1.00 |
| **Wheeze** | 11 (5%) | 2 (2%) | 8 (7%) | 1 (6%) | 0.21 | 8 (9%) | 15 (14%) | 3 (19%) | 0.44 | 8 (10%) | 20 (19%) | 3 (20%) | 0.16 |
| **Skin rash** | 5 (2%) | 2 (2%) | 3 (3%) | 0 (0%) | 1.00 | 5 (6%) | 4 (4%) | 0 (0%) | 0.76 | 8 (10%) | 10 (10%) | 0 (0%) | 0.60 |

Severe and critical groups combined due to low numbers of prospectively-enrolled critical patients (n=1). Groups compared using the Pearson $\chi^2$ test (or Fisher exact test if $n < 5$ ). Illness
onset defined as date of diagnosis for asymptomatic participants, who contributed to the denominator of the incidence proportion. No loss to follow-up occurred between day 1 and day 7
after illness onset. Loss to follow-up between day 7 and 28 after illness onset was minimal: n=3 in mild group, n=3 in moderate group, n=1 in severe group.

Transition plots showed that while most participants transitioned to a lower level of severity over time across the five persistent symptoms (fatigue, cough, dyspnoea, loss of smell and/or taste and myalgia), some transitioned to a higher severity level over time (Figure 1). Among those with mild disease, the majority of participants reported loss of smell and/or taste as severe at baseline, although most reported improvement within the first week of follow-up (Figures S2a-e).

**Figure 1a-e.**
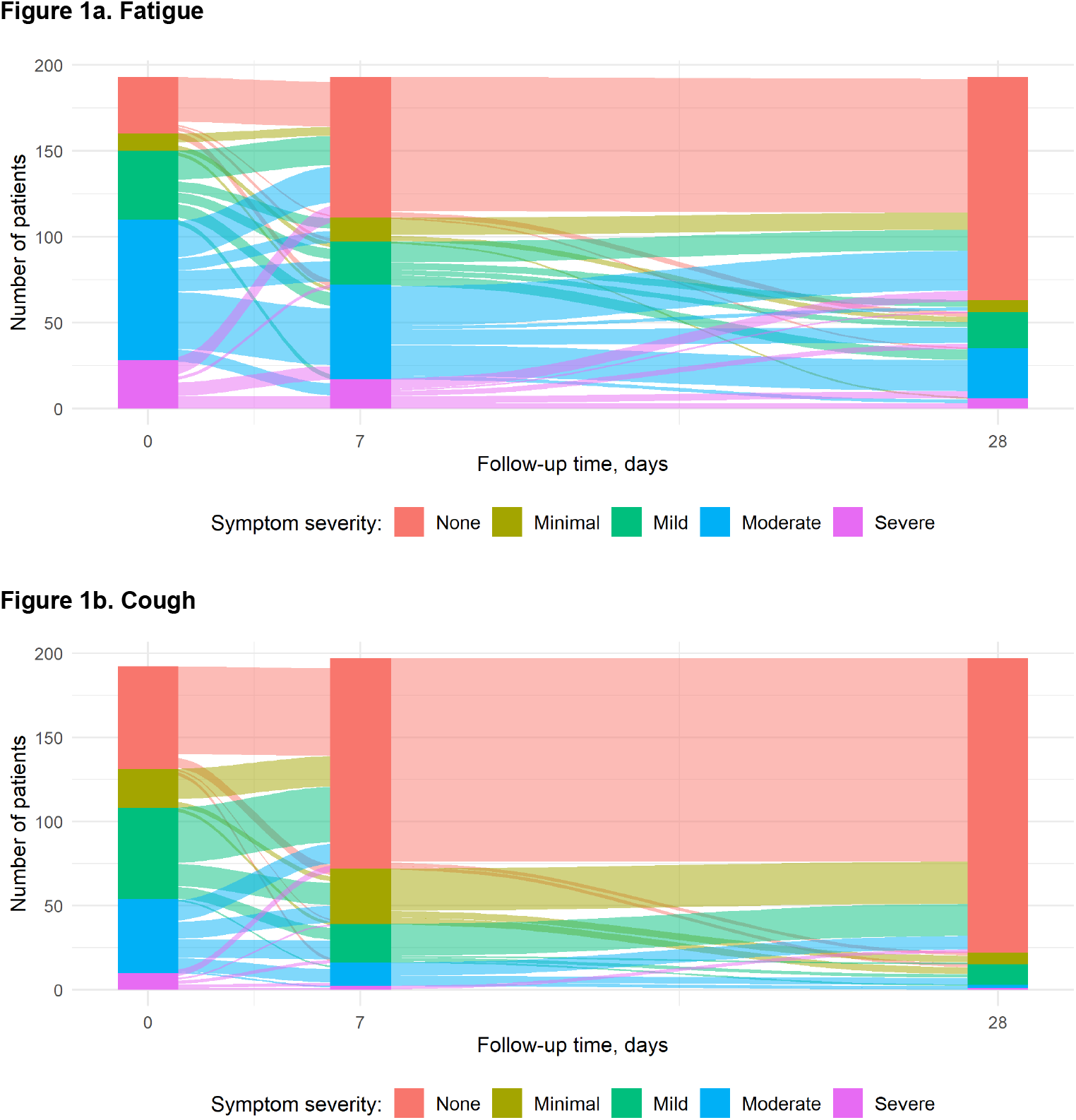
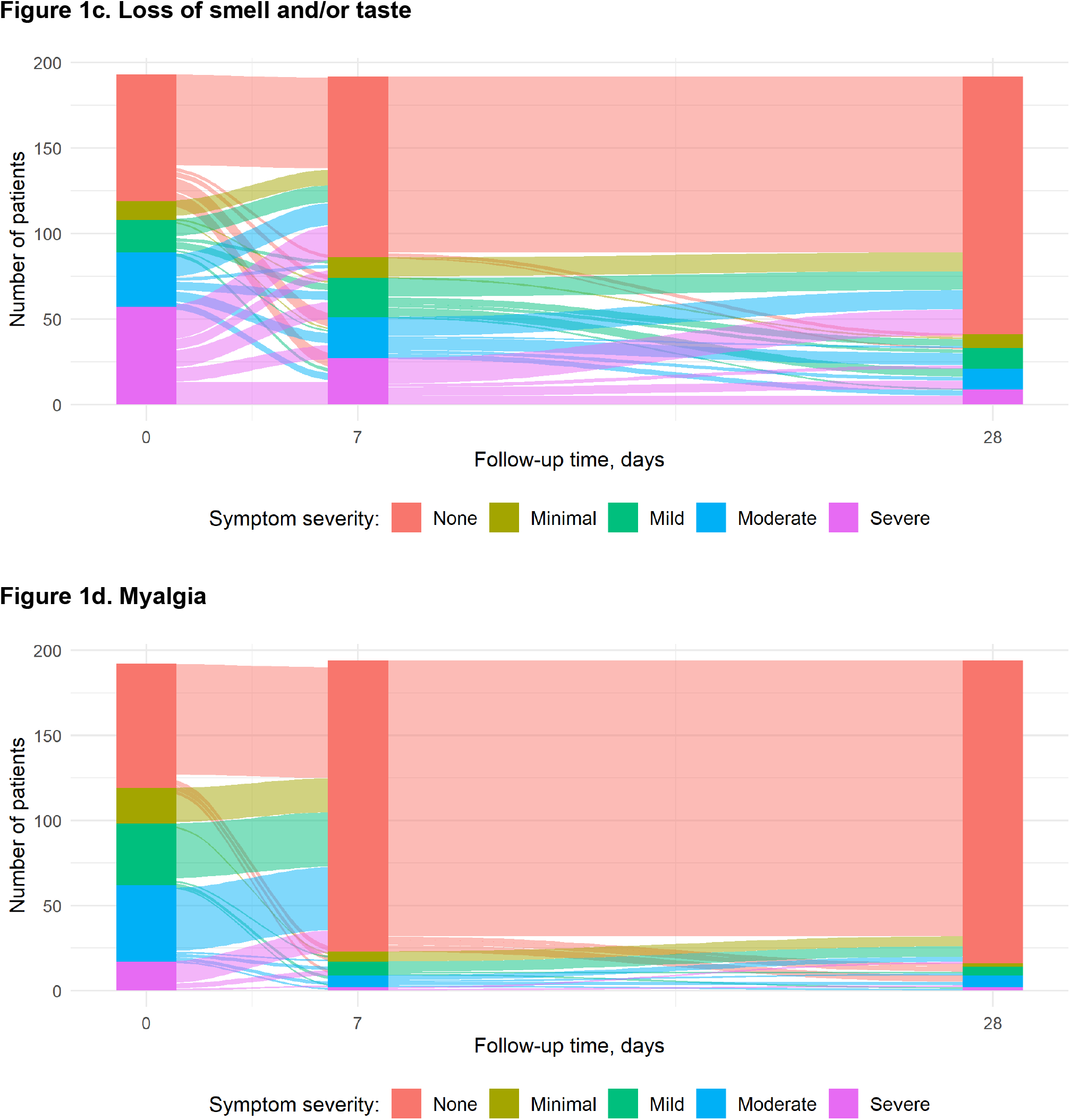
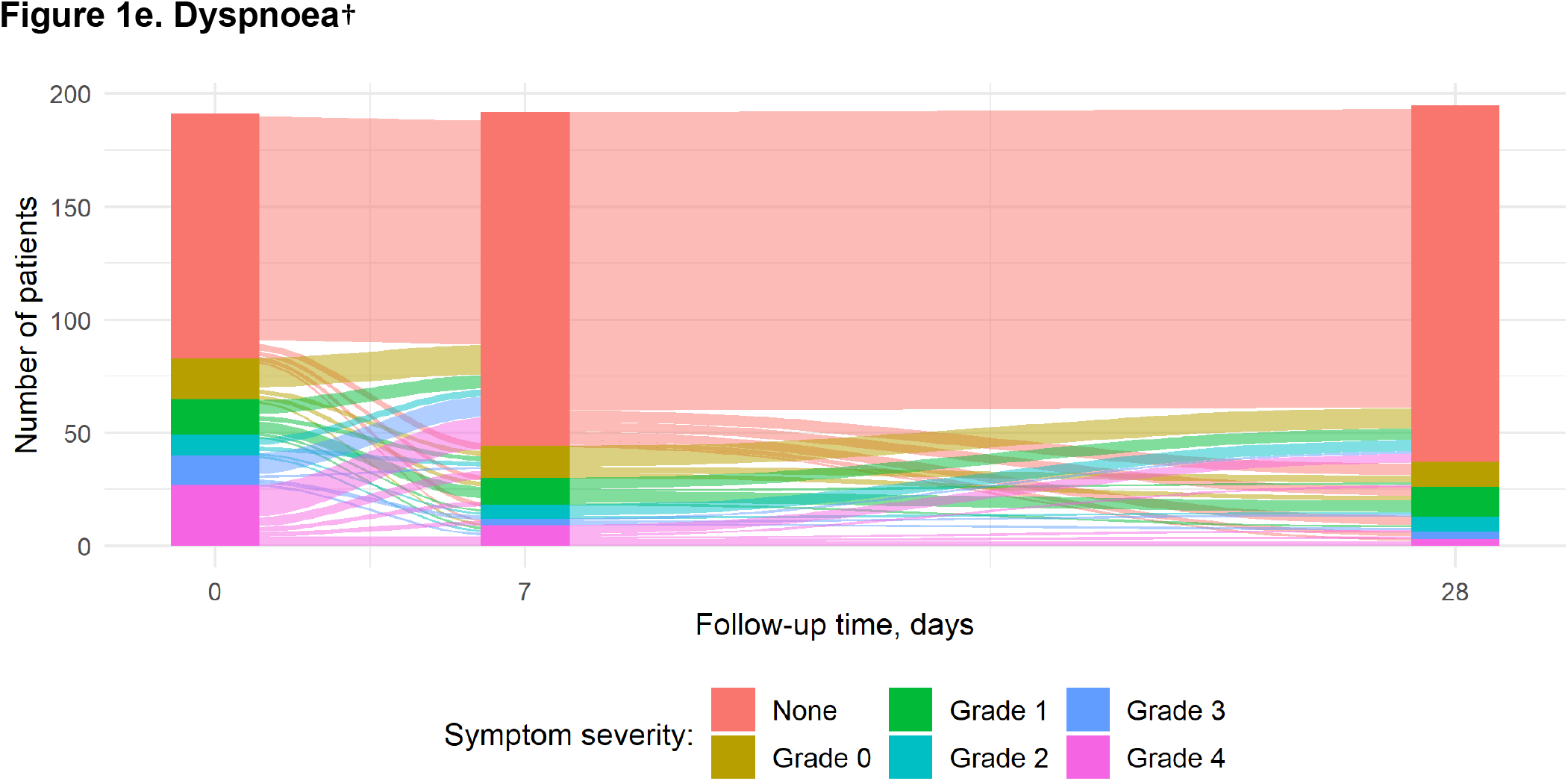
Transition between levels of self-reported presence and severity of fatigue, cough, loss of smell and/or taste, myalgia and dyspnoea over time among prospectively-included participants. Median time from illness onset to recruitment was 8 days (IQR=5-12), therefore visit days in transition plots approximately correspond to day 8, day 15 and day 36 since illness onset. Vertical bars represent the number of participants per severity level of the symptom at each day of follow-up. Severity level was defined as the maximum severity experienced by the study participant since the last study visit, i.e. the severity level reported at day 7 of follow-up represents the maximum severity experienced between recruitment and day 7 of follow-up. The size of the streams encodes the number of study participants who transitioned from one severity level to another; transitioning from any severity level to “none” represents recovery from that symptom. †mMRC Score: Grade 0 = breathlessness only on strenuous exercise; Grade 1 = shortness of breath when hurrying on the level or walking up a slight hill; Grade 2 = walks slower than people of same age on the level because of breathlessness or has to stop to catch breath when walking at their own pace on the level; Grade 3 = stops for breath after walking ∼100 m or after few minutes on the level; and Grade 4 = too breathless to leave the house, or breathless when dressing or undressing.

### Time to recovery from symptoms

Time to complete recovery was longer in symptomatic participants with moderate and severe/critical disease than in those with mild COVID-19 (log-rank p<0.001; Figure 2). At least one ongoing symptom was reported at 12 weeks after illness onset, by 33.0% (95%CI=23.0%-43.3%) of participants with mild, 63.9% (95%CI=54.8-71.6%) with moderate and 81.7% (95%CI=68.7-89.7%) with severe/critical disease. Among participants with mild disease, median time to complete recovery was 57 days (Figure 2), although 20.3%(95%CI=11.4-30.9%) continued to report at least one ongoing symptom at nine months after illness onset. Among participants with moderate and severe/critical disease, more than half reported at least one ongoing symptom at nine months after illness onset (53.0%[95%CI=43.2-61.8%] and 52.9%[95%CI=38.1-65.6%], respectively). Supplementary Figures 3a-e show Kaplan-Meier survival curves for the individual 18 symptoms, by clinical severity group.

**Figure 2.**
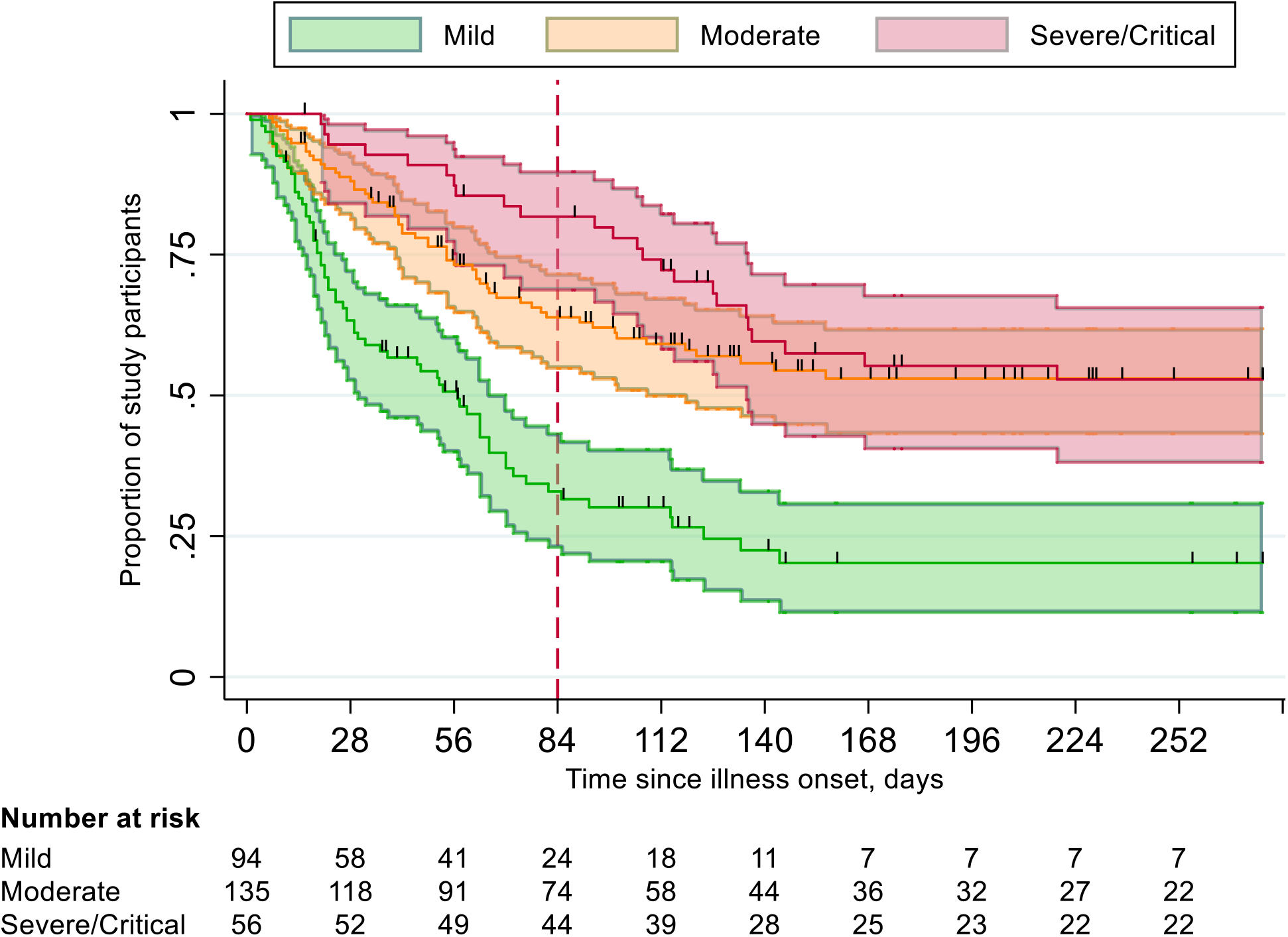
Kaplan-Meier plot of time from illness onset to complete recovery from symptoms, by clinical severity group. Clinical severity groups of severe and critically severe COVID-19 combined due to small numbers in the critical severity group. Dashed red line denotes cut-off point of 12 weeks (as per NICE definition of post-COVID syndrome); black vertical lines indicate time-points at which participants were censored.

**Figure 3.**
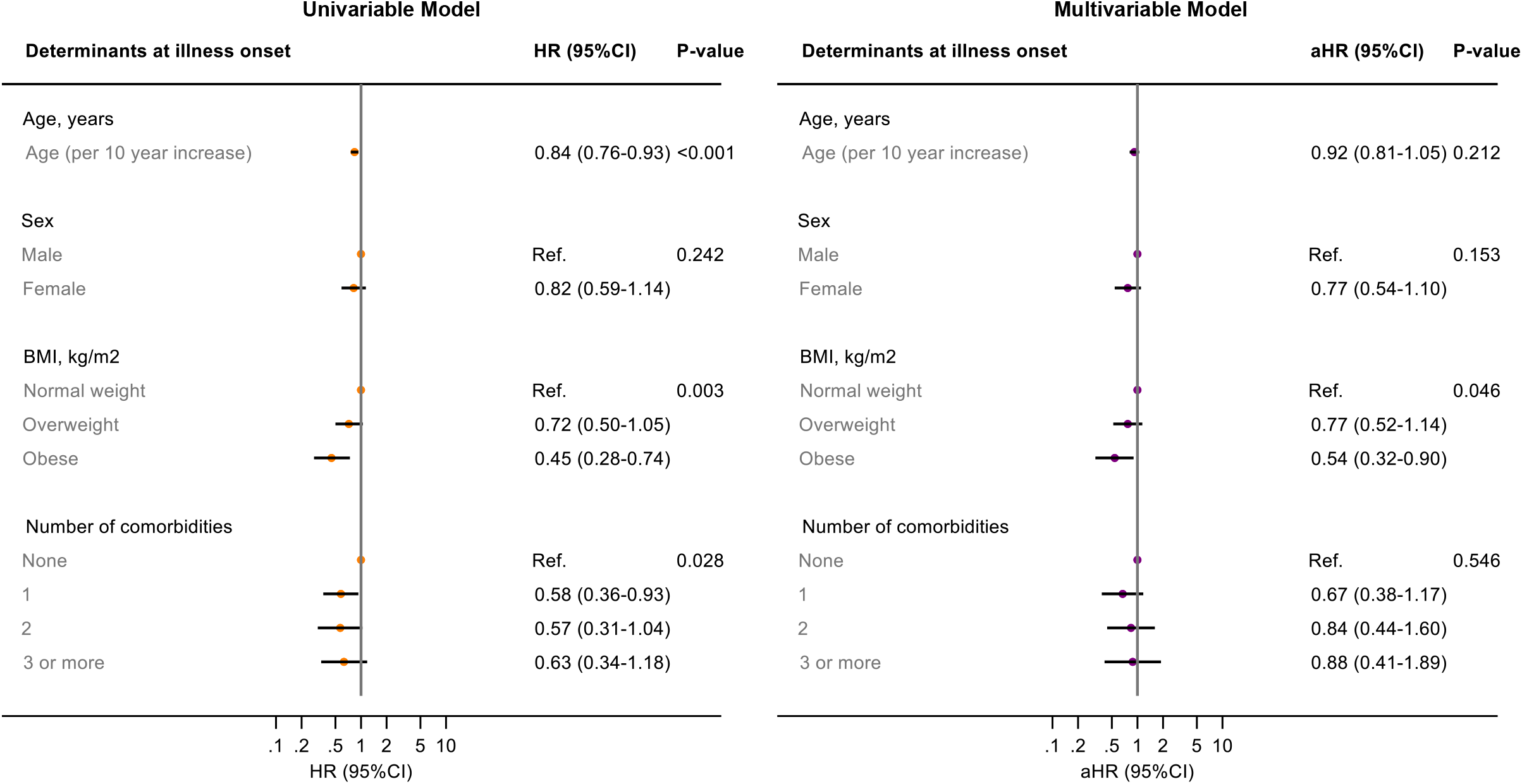
Univariable and multivariable Cox proportional hazard ratios of time to complete recovery for age, sex, BMI and comorbidity status at illness onset. Comorbidities counted are those listed by the WHO as being associated with a higher risk of developing severe or critical COVID-19[11]. Body mass index (BMI) categorised in kg/m^2^ as: <25, underweight or normal weight; 25 up to 30, overweight; >30, obese. P-value calculated using likelihood ratio test.

### Determinants of time to recovery from symptoms

BMI at illness onset showed the strongest association with time to complete recovery in multivariable analyses: obese participants had a 46% (aHR 0.54, 95%CI=0.32-0.90]) and overweight participants a 23% slower recovery (aHR 0.77, 95%CI=0.52-1.14]) compared to those of normal weight (p=0.046) (Figure 3). No statistically significant interactions were identified between BMI and the other covariates. Results were comparable in both sensitivity analyses (Figures S4a-b).

In multivariable analysis of time to recovery from each of the five most persistent symptoms (Table S1a-e), being obese at illness onset was associated with slower recovery from cough (aHR 0.56, 95%CI=0.35-0.89) and loss of smell and/or taste (aHR 0.44, 95%CI=0.25-0.76). Increased age was associated with slower recovery from cough (aHR 0.81, 95%CI=0.72-0.92), dyspnoea (aHR 0.82, 95%CI=0.70-0.96), and myalgia (aHR 0.74, 95%CI=0.65-0.85). Number of comorbidities at illness onset was only associated with recovery from fatigue, where those with one comorbidity recovered more than twice as slowly as those without comorbidities (aHR 0.45, 95%CI=0.28-0.72). When we replaced number of comorbidities with the presence of each of CVD, CLD, DM or immunodeficiency in the multivariable models for each of these symptoms, no statistically significant effect on time to recovery was detected for any of these specific comorbidities.

## Discussion

To our knowledge, this study is the first to report detailed longitudinal data on the evolution of COVID-19 symptoms in a cohort of individuals with mild to critical disease. We observed that the incidence of certain hallmark COVID-19 symptoms were comparable between clinical severity groups during the acute phase of illness. Despite an overall improvement in severity of the most persistent COVID-19 symptoms during the first month of illness, one-third of the mild group, nearly two-thirds of the moderate group and more than four-fifths of patients with severe/critical disease met NICE criteria for post-COVID syndrome. Even at nine months after illness onset, one-fifth of those with mild disease and over half of participants with either moderate or severe/critical disease experienced at least one ongoing symptom. Obesity at illness onset was the strongest determinant of slow recovery from symptoms.

Since the start of the COVID-19 pandemic, avoiding the immediate consequences of hospitalisation and mortality has been the primary goal. Longer term sequelae of COVID-19 have received relatively little attention, especially among non-hospitalised patients. In our study, the proportion of participants reporting fatigue, fever and cough during the first month of illness were as high in mild COVID-19 patients as they were in moderate, severe or critical patients. Although time to recovery from symptoms was shorter in those with mild disease, as many as one in three participants with mild COVID-19 still reported symptoms 12 weeks after illness onset. Indeed, the proportion of participants meeting the NICE definition of post-COVID syndrome in our cohort (57.8% overall) was comparable to other prospective cohort studies[15, 16], but higher than estimates by the UK Office for National Statistics and among healthcare workers [17, 18]. Although this could be partly explained by the fact that we limited our analysis to symptomatic participants, the consequences of these proportions when extrapolated to a global level may be substantial. As new viral variants and challenges in vaccination programmes worldwide continue to hinder effective control measures, the number of people with post-COVID syndrome will only continue to increase.

Although patient advocacy groups have helped in making post-COVID syndrome a research priority[19], studies to date have differed in study population, follow-up time and symptoms evaluated [9], making it difficult to synthesize all available evidence. Moreover, the symptom profiles that falls under post-COVID syndrome are diverse [20], resulting in a heterogenous patient group requiring different management strategies. A universally accepted and evidence-based definition of post-COVID syndrome is key to comparing findings across studies and settings, and to develop syndrome-specific interventions. Moreover, an validated risk score for post-COVID syndrome could help prevention and identification of those requiring support. Our study, for example, suggests that obese individuals, regardless of sex, age and number of comorbidities at illness onset, may benefit from early intervention. In addition to its independent effect on recovery, obesity is associated with having a lower socio-economic status and reduced access to health and care services[21], both of which may also be associated with a slower recovery from symptoms. Over and above the impact of on acute COVID-19 hospitalisation and mortality [4, 22], reducing the prevalence of obesity may also help reduce the future burden of post-COVID syndrome.

Fatigue was the most commonly reported symptom both during the acute phase and at 12 weeks from illness onset, including among individuals with mild or moderate disease. Previous analyses have estimated that the societal impact of fatigue can be significant, due to both direct healthcare costs and indirect financial losses resulting from reduced economic productivity[23]. As those with mild COVID-19 represent the majority of COVID-19 cases worldwide in terms of absolute numbers, developing strategies to prevent, diagnose and manage post-COVID fatigue should be an urgent research priority. Among participants with moderate and severe/critical disease, dyspnoea and myalgia additionally persisted beyond 12 weeks in a large proportion of participants. Similar results have been reported in other settings: previously-hospitalised COVID-19 patients in Wuhan, China still had abnormal chest imaging findings and pulmonary diffusing capacity at 6 months after illness onset [7], whilst a cross-sectional study of hospitalised COVID-19 patients in the UK reported that the majority of participants reported myalgia at a median follow-up of 16 weeks after discharge from hospital[24]. In our multivariable analysis, older age was the most important determinant of slower time to recovery from both of these symptoms. Exploring the underlying mechanism as to why these symptoms persist in older patients may help identify interventions that could be beneficial in the recovery process.

This study has several strengths. Frequent symptom questionnaires collected longitudinally since illness onset allowed the natural progression of COVID-19 symptoms to be described to a level of detail not previously reported. We were able to enrol patients with mild symptoms (underrepresented in other studies) as well as those who were critically ill, so that the full spectrum of COVID-19 disease could be represented. Several limitations must be recognised. Survival bias among the retrospectively-enrolled hospitalised participants may have caused an underestimation of time to complete recovery in those with severe/critical disease, although results were comparable when restricting our analyses to prospectively identified participants. Questionnaires in languages other than Dutch and English were not offered, therefore individuals with a migration background, who have been disproportionally affected by COVID-19, also in Amsterdam[25, 26], were underrepresented in this cohort. Furthermore, misclassification bias may have resulted from using ICU admission as a proxy for critical disease; suitability for ICU admission is also judged by the patient’s chance of survival; indeed, those with critical disease tended to be younger and have a lower BMI than those in the severe group. Finally, the natural progression of disease may differ between SARS-CoV-2 variants[27] and treatments received. Therefore, our results may not be representative for future patients infected with SARS-CoV-2, especially as the treatment landscape for COVID-19 changes and new variants continue to arise.

We demonstrated that post-COVID syndrome is common, even after mild disease. Symptoms persisted for nine months after illness onset in one-fifth of participants with mild disease and in more than half of participants with moderate and severe/critical disease. Obesity was the most important predictor of slow recovery and thus creating an environment which facilitates healthy living behaviours is of utmost importance, even during a pandemic. Next steps in post-COVID syndrome research must include assessing the public health and socioeconomic impact, identifying further predictive and prognostic characteristics, and exploring the underlying biological mechanisms of disease in order to identify and develop effective interventions.

### RECoVERED Study Group

Public Health Service of Amsterdam: Ivette Agard, Jane Ayal, Floor Cavder, Marianne Craanen, Udi Davidovich, Annemarieke Deuring, Annelies van Dijk, Ertan Ersan, Laura del Grande, Joost Hartman, Nelleke Koedoet, Romy Lebbink, Dominique Loomans, Agata Makowska, Tom du Maine, Ilja de Man, Lizenka van der Meij, Marleen van Polanen, Maria Oud, Clark Reid, Leeann Storey, Marc van Wijk Amsterdam University Medical Centres: Joyce van Assem, Marijne van Beek, Eric Moll van Charante, Orlane Figaroa, Leah Frenkel, Xiaochuan (Alvin) Han, Agnes Harskamp-Holwerda, Mette Hazenberg, Soemeja Hidad, Nina de Jong, Hans Knoop, Lara Kuijt, Anja Lok, Pythia Nieuwkerk, Colin Russell, Karlijn van der Straten, Annelou van der Veen, Bas Verkaik, Anouk Verveen, Gerben-Rienk Visser

## Data Availability

Information can be obtained from the corresponding author, Elke Wynberg

## Acknowledgements

The authors wish to thank all RECoVERED study participants. Additionally we to thank Daniëla van Santen for helping to create the transition plots.

## Notes

### Competing Interest Statement

The authors have declared no competing interest.

### Funding Statement

The RECoVERED study received funding from ZonMw (project number 10150062010002) and a Research & Development grant (21-14) from the Public Health Service of Amsterdam

### Author Declarations

Medical Ethics Examination Committee (METC) of the Amsterdam Medical Centers

