## Supplementary Tables & Figures for "Evolution of COVID-19 symptoms during the first 9 months after illness onset"

### Supplementary Files

**Supplementary Table 1a-e.** Association between age, sex, BMI and number of comorbidities at illness onset and time to recovery from fatigue, cough, dyspnoea, loss of smell and/or taste and myalgia in univariable and multivariable Cox proportional hazard models

**Supplementary Figure 1.** Overview of data collection in the RECoVERED Study, Amsterdam, the Netherlands

**Supplementary Figures 2a-e.** Transition between levels of self-reported severity of (a) fatigue, (b) cough, (c) loss of smell and/or taste, (d) myalgia and (e) dyspnoea over time among prospectively-included participants, by clinical severity group

**Supplementary Figures 3a-e.** Kaplan-Meier plots of time to recovery from each individual COVID-19 symptom, by clinical severity group

**Supplementary Figures 4a-b.** Sensitivity analyses: Multivariable Cox proportional hazards models of time from illness onset to complete recovery when (a) restricting to prospectively-included participants only and (b) left-truncating data at date of enrolment into the study

**Table S1a-e. Association between age, sex, BMI and number of comorbidities at illness onset and time to recovery from fatigue, cough, dyspnoea, loss of smell and/or taste and myalgia in univariable and multivariable Cox proportional hazard models**

| | Univariable model | | | | Multivariable model | | | |
|---|---|---|---|---|---|---|---|---|
| **Table 3a. Fatigue** | | | | | | | | |
| **Baseline variable** | **HR** | **95% CI** | | **p-value** | **HR** | **95% CI** | | **p-value** |
| Age, years (per 10 year increase)* | 0.826 | 0.747 | 0.912 | <0.001 | 1.003 | 1.001 | 1.005 | 0.010 |
| Sex | | | | 0.842 | | | | 0.614 |
| Male | Ref. | | | | Ref. | | | |
| Female | 0.970 | 0.718 | 1.311 | | 0.920 | 0.667 | 1.270 | |
| BMI | | | | 0.038 | | | | 0.101 |
| Normal weight | Ref. | | | | Ref. | | | |
| Overweight | 0.739 | 0.521 | 1.047 | | 0.797 | 0.555 | 1.145 | |
| Obese | 0.599 | 0.392 | 0.914 | | 0.627 | 0.401 | 0.979 | |
| Number of comorbidities | | | | 0.005 | | | | 0.003 |
| 0 | Ref. | | | | Ref. | | | |
| 1 | 0.529 | 0.349 | 0.802 | | 0.450 | 0.283 | 0.717 | |
| 2 | 0.789 | 0.479 | 1.299 | | 0.923 | 0.546 | 1.559 | |
| 3 or more | 0.562 | 0.322 | 0.980 | | 0.556 | 0.288 | 1.073 | |
| **Table 3b. Cough** | | | | | | | | |
| **Baseline variable** | **HR** | **95% CI** | | **p-value** | **HR** | **95% CI** | | **p-value** |
| Age, years (per 10 year increase) | 0.810 | 0.733 | 0.896 | <0.001 | 0.810 | 0.716 | 0.917 | <0.001 |
| Sex | | | | 0.344 | | | | 0.247 |
| Male | Ref. | | | | Ref. | | | |
| Female | 1.160 | 0.854 | 1.576 | | 1.213 | 0.876 | 1.680 | |
| BMI | | | | 0.004 | | | | 0.036 |
| Normal weight | Ref. | | | | Ref. | | | |
| Overweight | 0.767 | 0.539 | 1.091 | | 0.756 | 0.528 | 1.084 | |
| Obese | 0.492 | 0.317 | 0.765 | | 0.559 | 0.350 | 0.893 | |
| Number of comorbidities | | | | 0.005 | | | | 0.690 |
| 0 | Ref. | | | | Ref. | | | |
| 1 | 0.639 | 0.421 | 0.970 | | 0.833 | 0.516 | 1.345 | |
| 2 | 0.693 | 0.415 | 1.160 | | 1.151 | 0.647 | 2.049 | |
| 3 or more | 0.416 | 0.228 | 0.759 | | 0.785 | 0.384 | 1.606 | |
| **Table 3c. Dyspnoea** | | | | | | | | |
| **Baseline variable** | **HR** | **95% CI** | | **p-value** | **HR** | **95% CI** | | **p-value** |
| Age, years (per 10 year increase) | 0.741 | 0.651 | 0.842 | <0.001 | 0.820 | 0.697 | 0.962 | 0.013 |
| Sex | | | | 0.760 | | | | 0.373 |
| Male | Ref. | | | | Ref. | | | |
| Female | 0.943 | 0.647 | 1.374 | | 0.831 | 0.551 | 1.251 | |
| BMI | | | | 0.008 | | | | 0.102 |
| Normal weight | Ref. | | | | Ref. | | | |
| Overweight | 0.743 | 0.487 | 1.133 | | 0.795 | 0.508 | 1.243 | |
| Obese | 0.421 | 0.237 | 0.747 | | 0.526 | 0.286 | 0.968 | |
| Number of comorbidities | | | | 0.003 | | | | 0.492 |
| 0 | Ref. | | | | Ref. | | | |
| 1 | 0.435 | 0.253 | 0.748 | | 0.649 | 0.349 | 1.205 | |
| 2 | 0.527 | 0.286 | 0.970 | | 0.842 | 0.434 | 1.632 | |
| 3 or more | 0.552 | 0.285 | 1.070 | | 1.055 | 0.479 | 2.320 | |

| **Table 3d. Myalgia** | | | | | | | | |
|---|---|---|---|---|---|---|---|---|
| **Baseline variable** | **HR** | **95% CI** | | **p-value** | **HR** | **95% CI** | | **p-value** |
| Age, years (per 10 year increase) | 0.730 | 0.650 | 0.820 | <0.001 | 0.742 | 0.646 | 0.851 | <0.001 |
| Sex | | | | 0.473 | | | | 0.659 |
| Male | Ref. | | | | Ref. | | | |
| Female | 1.124 | 0.818 | 1.544 | | 1.084 | 0.759 | 1.549 | |
| BMI | | | | 0.083 | | | | 0.261 |
| Normal weight | Ref. | | | | Ref. | | | |
| Overweight | 0.842 | 0.580 | 1.223 | | 0.858 | 0.580 | 1.269 | |
| Obese | 0.600 | 0.378 | 0.953 | | 0.677 | 0.420 | 1.092 | |
| Number of comorbidities | | | | 0.004 | | | | 0.565 |
| 0 | Ref. | | | | Ref. | | | |
| 1 | 0.567 | 0.372 | 0.864 | | 0.716 | 0.439 | 1.167 | |
| 2 | 0.573 | 0.338 | 0.973 | | 0.885 | 0.486 | 1.610 | |
| 3 or more | 0.442 | 0.214 | 0.910 | | 0.733 | 0.318 | 1.687 | |
| **Table 3e. Loss of smell and/or taste** | | | | | | | | |
| **Baseline variable** | **HR** | **95% CI** | | **p-value** | **HR** | **95% CI** | | **p-value** |
| Age, years (per 10 year increase) | 0.930 | 0.842 | 1.027 | 0.151 | 0.934 | 0.823 | 1.061 | 0.292 |
| Sex | | | | 0.127 | | | | 0.267 |
| Male | Ref. | | | | Ref. | | | |
| Female | 0.780 | 0.566 | 1.074 | | 0.825 | 0.587 | 1.159 | |
| BMI | | | | 0.010 | | | | 0.008 |
| Normal weight | Ref. | | | | Ref. | | | |
| Overweight | 0.847 | 0.588 | 1.220 | | 0.791 | 0.538 | 1.163 | |
| Obese | 0.491 | 0.301 | 0.801 | | 0.437 | 0.253 | 0.756 | |
| Number of comorbidities | | | | 0.406 | | | | 0.213 |
| 0 | Ref. | | | | Ref. | | | |
| 1 | 0.736 | 0.475 | 1.142 | | 0.944 | 0.554 | 1.611 | |
| 2 | 0.864 | 0.502 | 1.489 | | 1.529 | 0.818 | 2.857 | |
| 3 or more | 1.107 | 0.578 | 2.120 | | 2.020 | 0.910 | 4.490 | |

Comorbidities counted are those listed by the WHO as being associated with a higher risk of developing severe or critical COVID-19[11]. Body mass index (BMI) categorised in kg/m$^2$ as: <25, underweight or normal weight; 25 up to 30, overweight; >30, obese.

*Age included as a time-varying covariate in the multivariable model of time to recovery from fatigue, to ensure the proportional hazards assumption was met.

**Figure S1. Overview of data collection in the RECoVERED Study, Amsterdam, the Netherlands.**

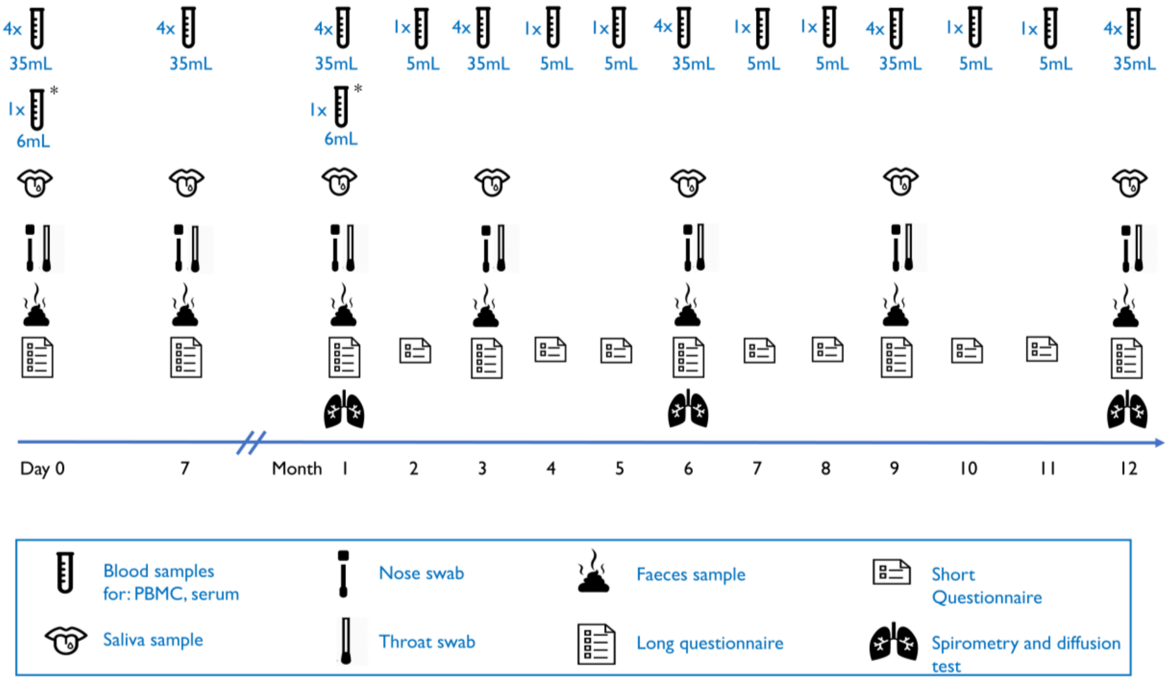


Long questionnaire: Socio-demographic, clinical, symptom and psychosocial data. Short questionnaire: Symptom data only.

**Figures S2a-e. Transition between levels of self-reported severity of (a) fatigue, (b) cough, (c) loss of smell and/or taste, (d) myalgia and (e) dyspnoea over time among prospectively-included participants, by clinical severity group**

Median time from illness onset to recruitment was 8 days (IQR 5-11), therefore day 0, day 7 and day 28 of follow-up in transition plots approximately correspond to day 8, day 15 and day 36 of natural disease progression. Vertical bars represent the number of participants per severity level of the symptom at each day of follow-up. Severity level was defined as the maximum severity experienced by the study participant since the last study visit, i.e. the severity level reported at day 7 of follow-up represents the maximum severity experienced between day 0 and 7 of follow-up. The size of the streams encodes the number of study participants who transitioned from one severity level to another; transitioning from any severity level to "none" represents recovery from that symptom.

**Figure S2a. Fatigue**

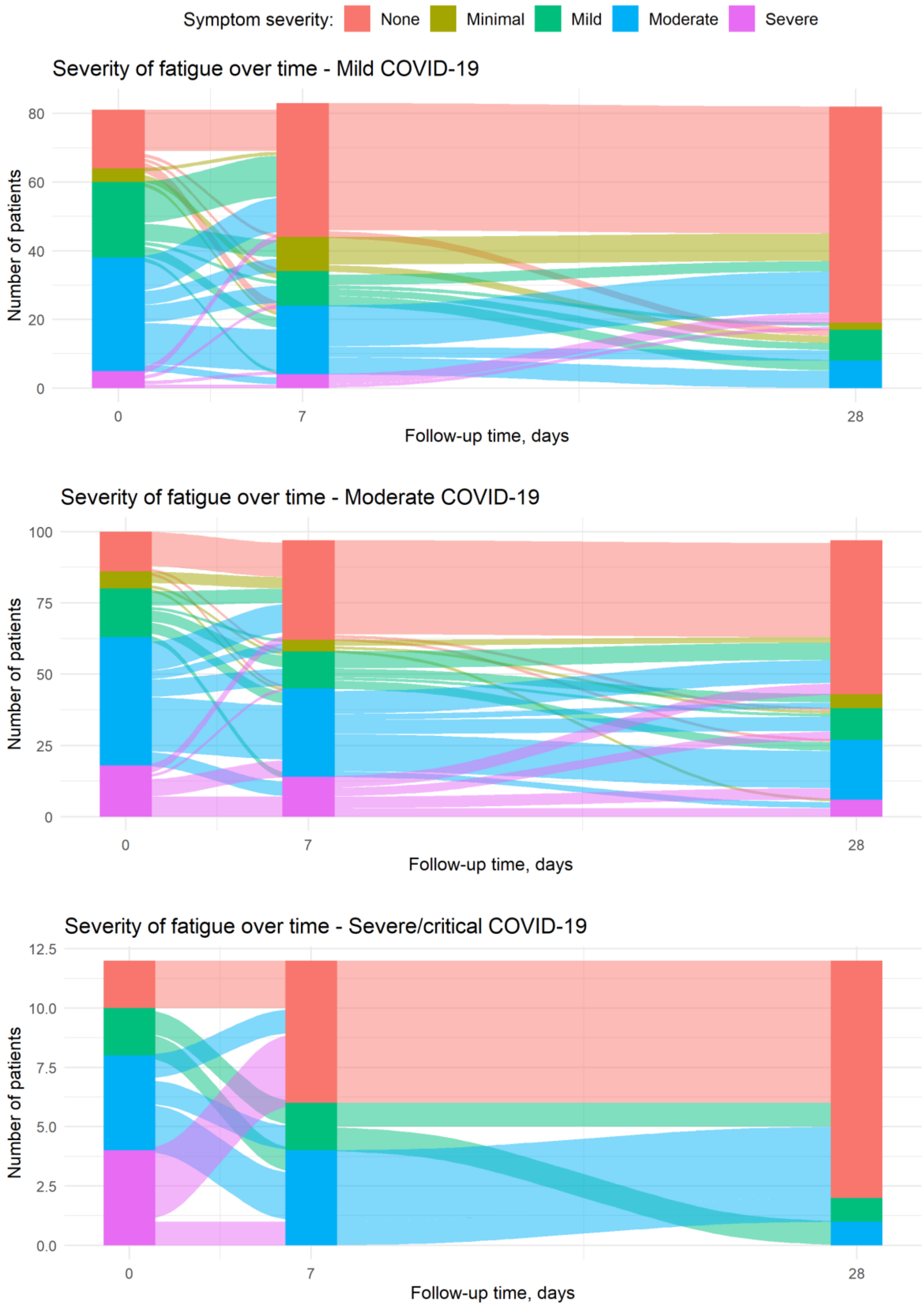

**Figure S2b. Cough**

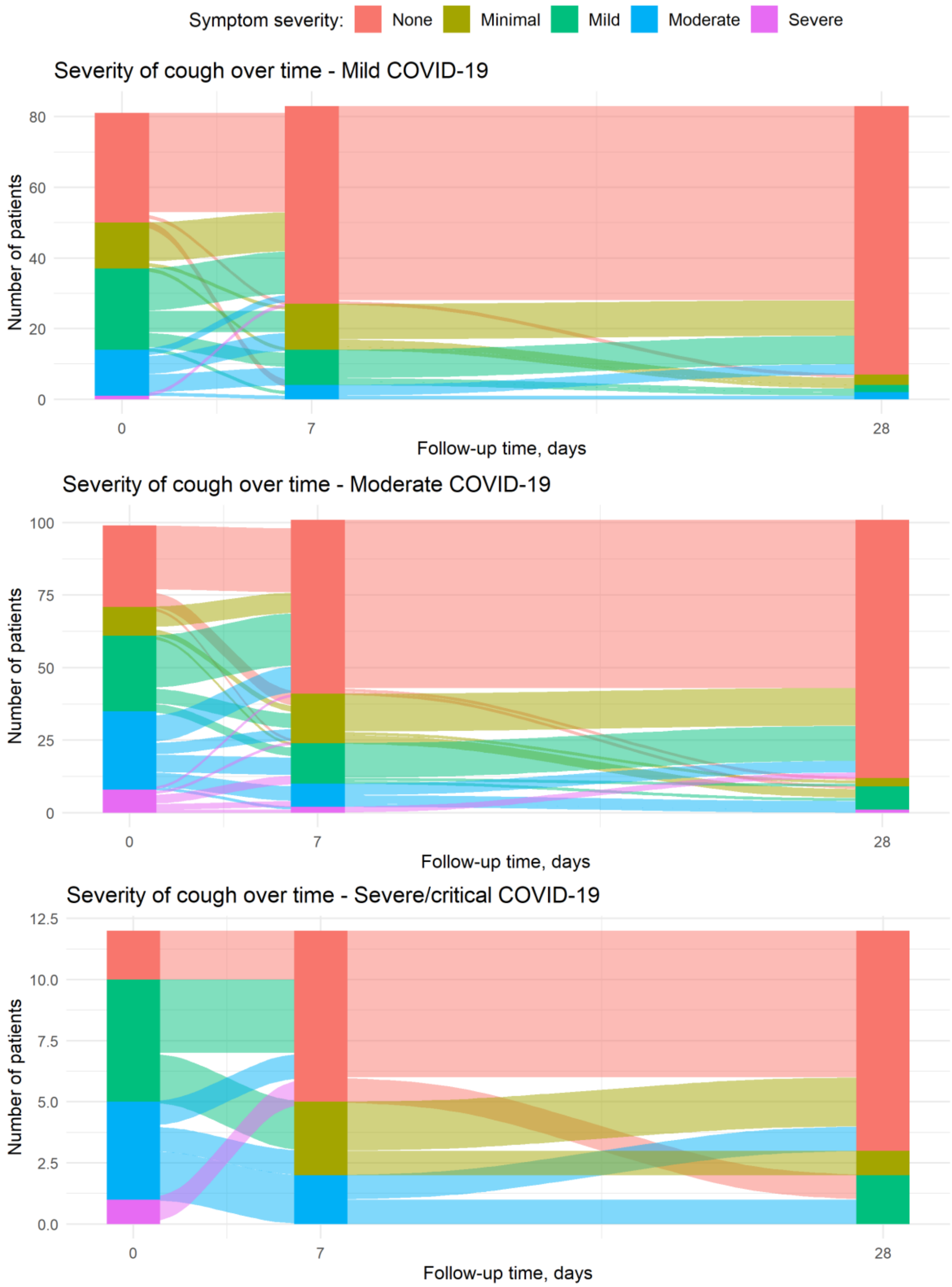

**Figure S2c. Loss of smell and/or taste**

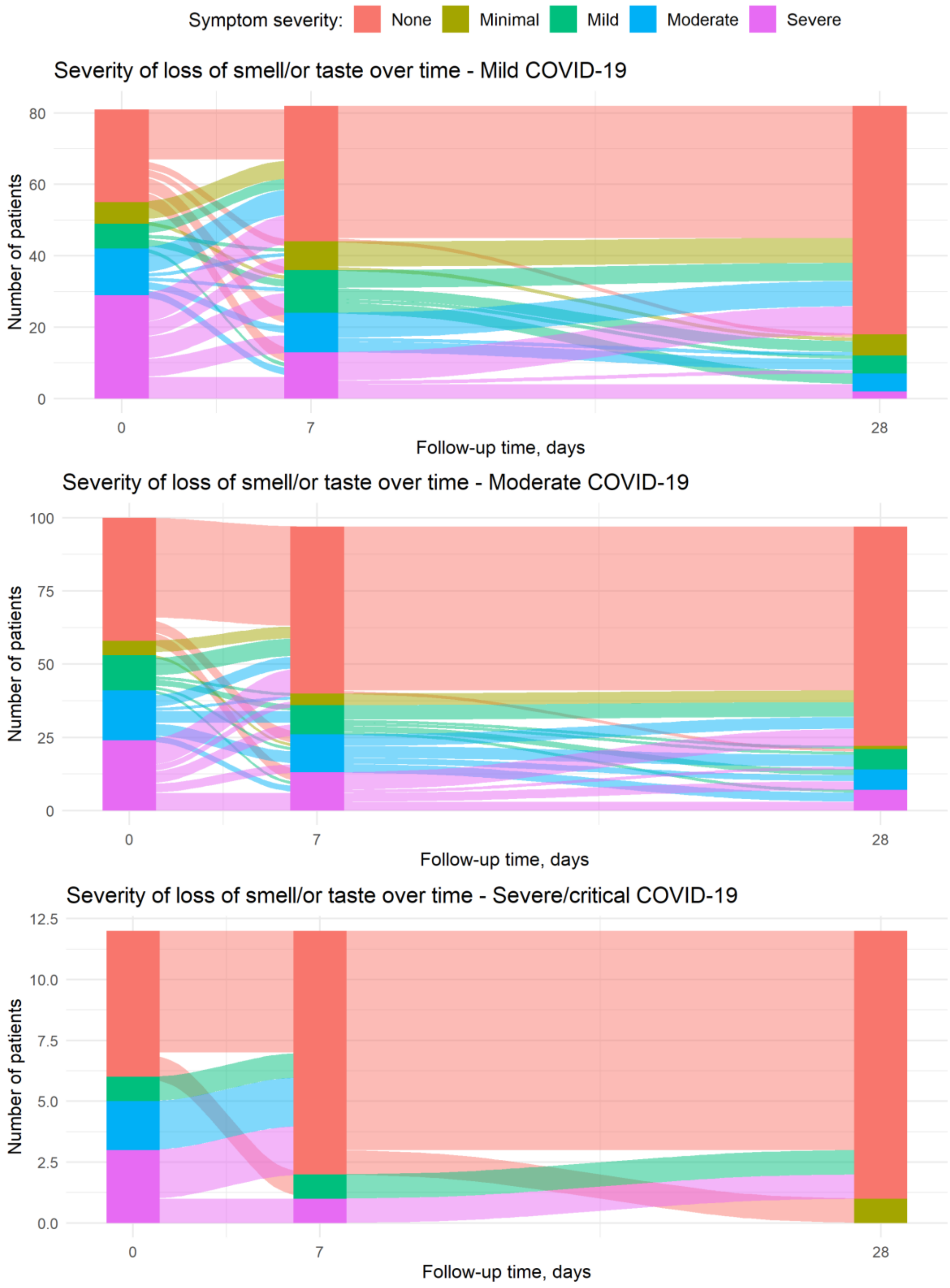

**Figure S2d. Myalgia**

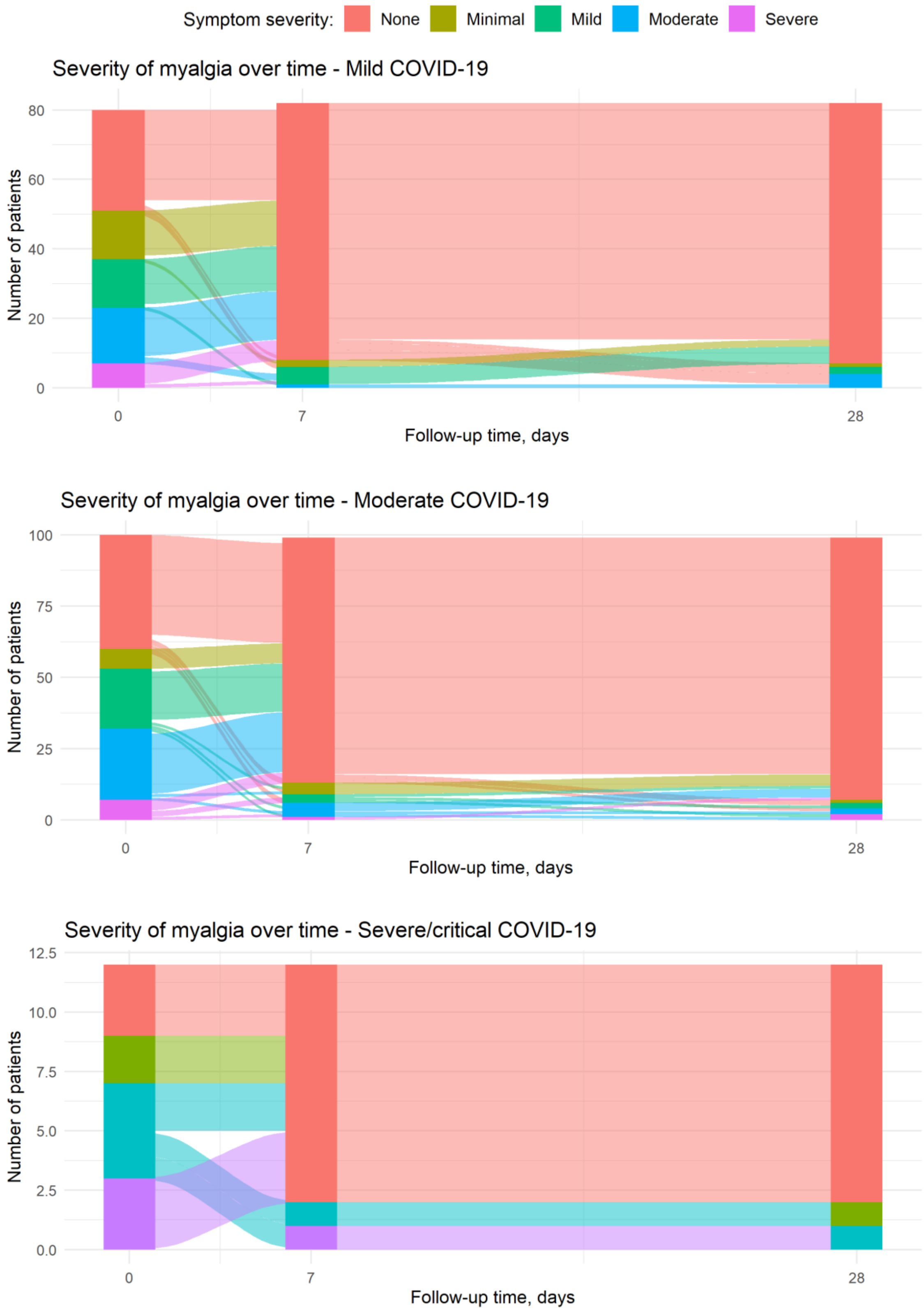

**Figure S2e. Dyspnoea**

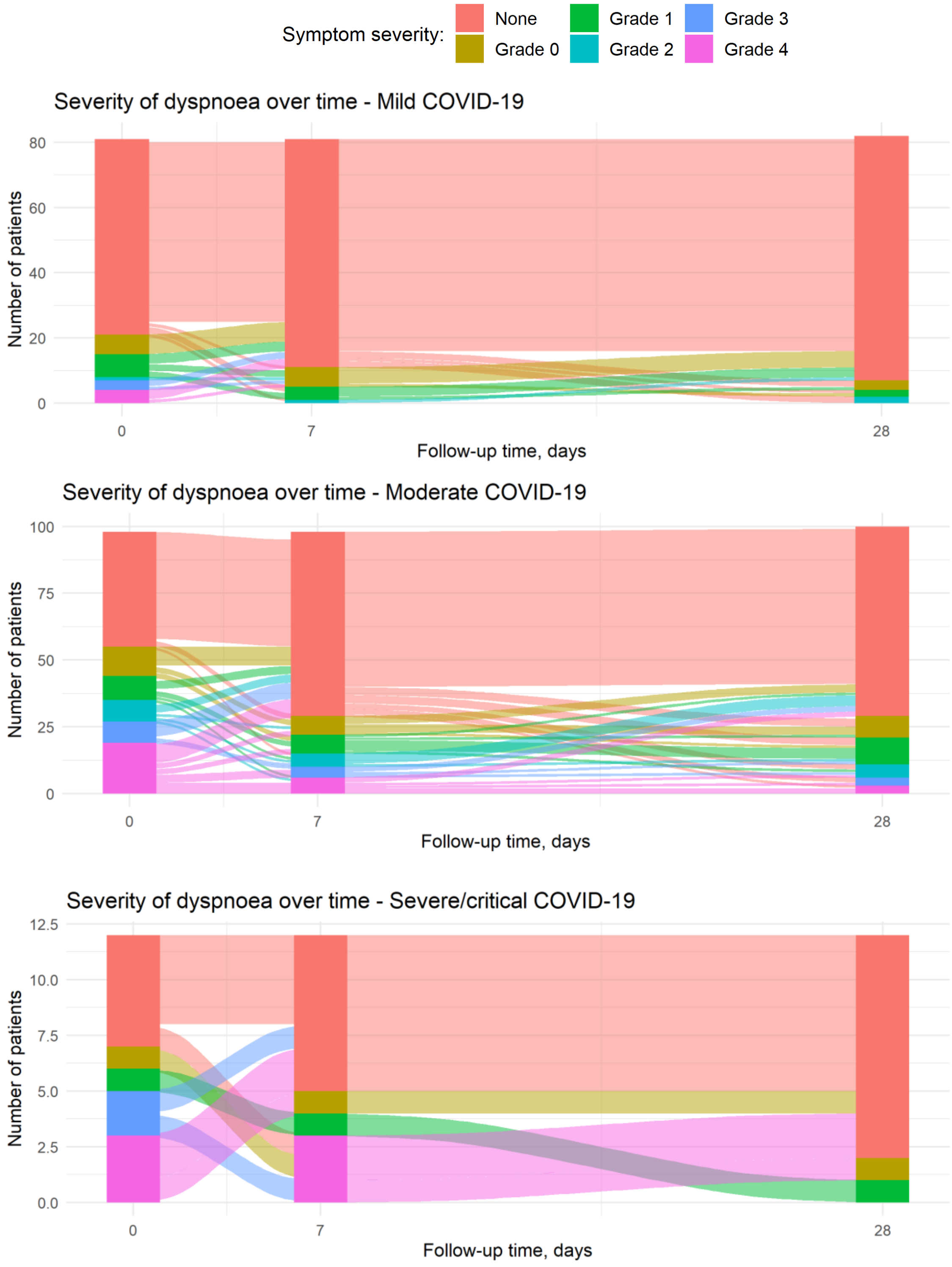

#### Figures S3a-e. Kaplan-Meier plots of time to recovery from each individual COVID-19 symptom, by clinical severity group

Red dotted line denotes 12 weeks post-illness onset (NICE criterion for post-COVID syndrome). 18 individual symptoms presented.

**Figure S3a. Fatigue, fever, cough and dyspnoea**

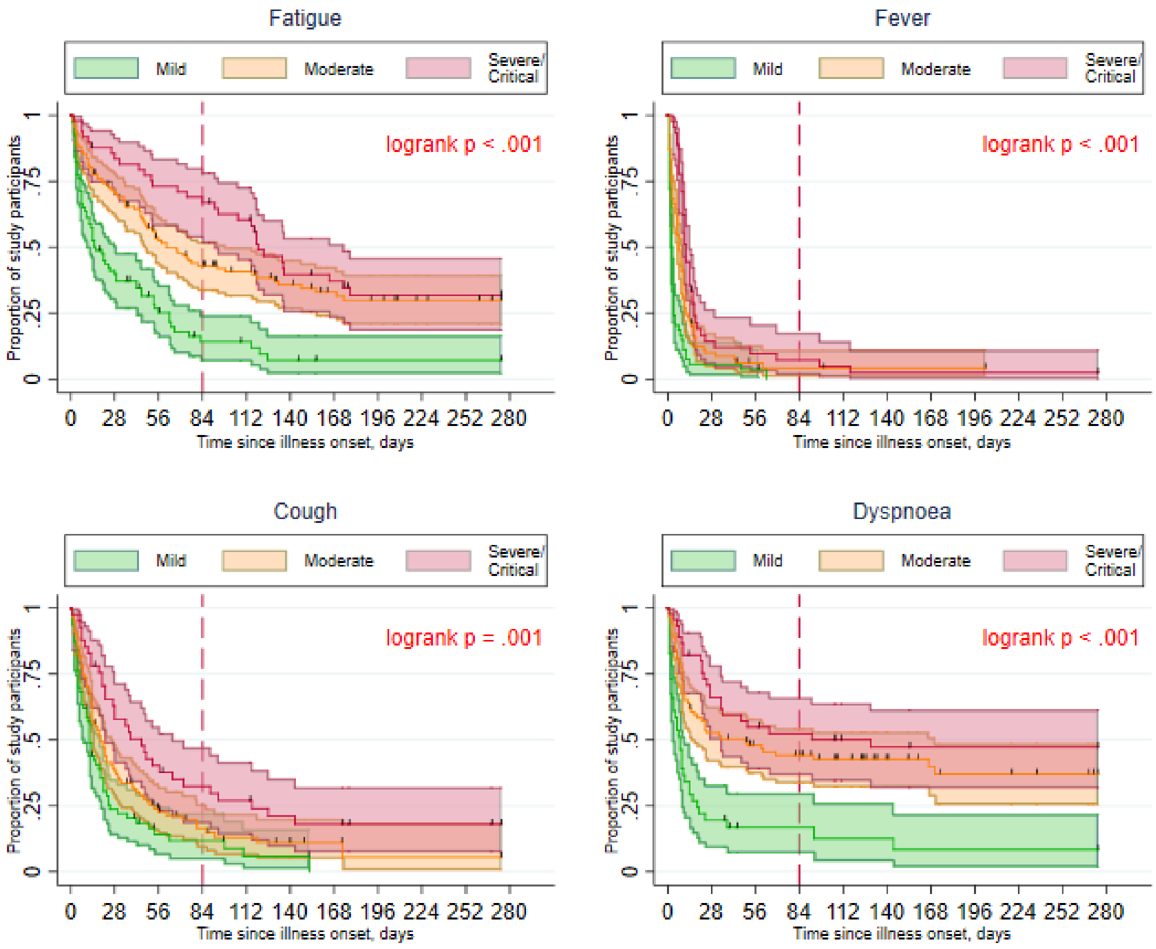

**Figure S3b. Loss of smell and taste, myalgia, headache and loss of appetite**

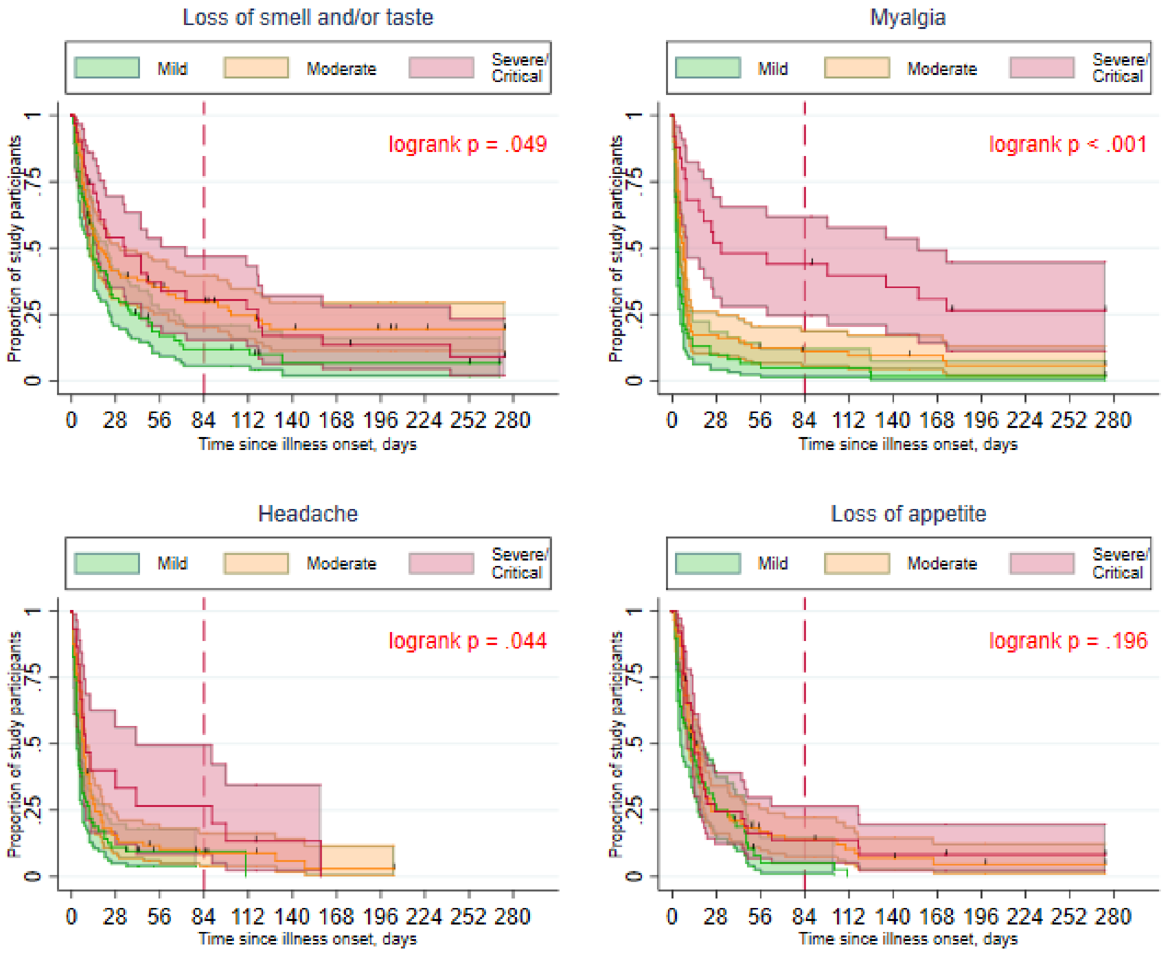

**Figure S3c. Rhinorrhoea, sore throat, wheeze and chest pain**

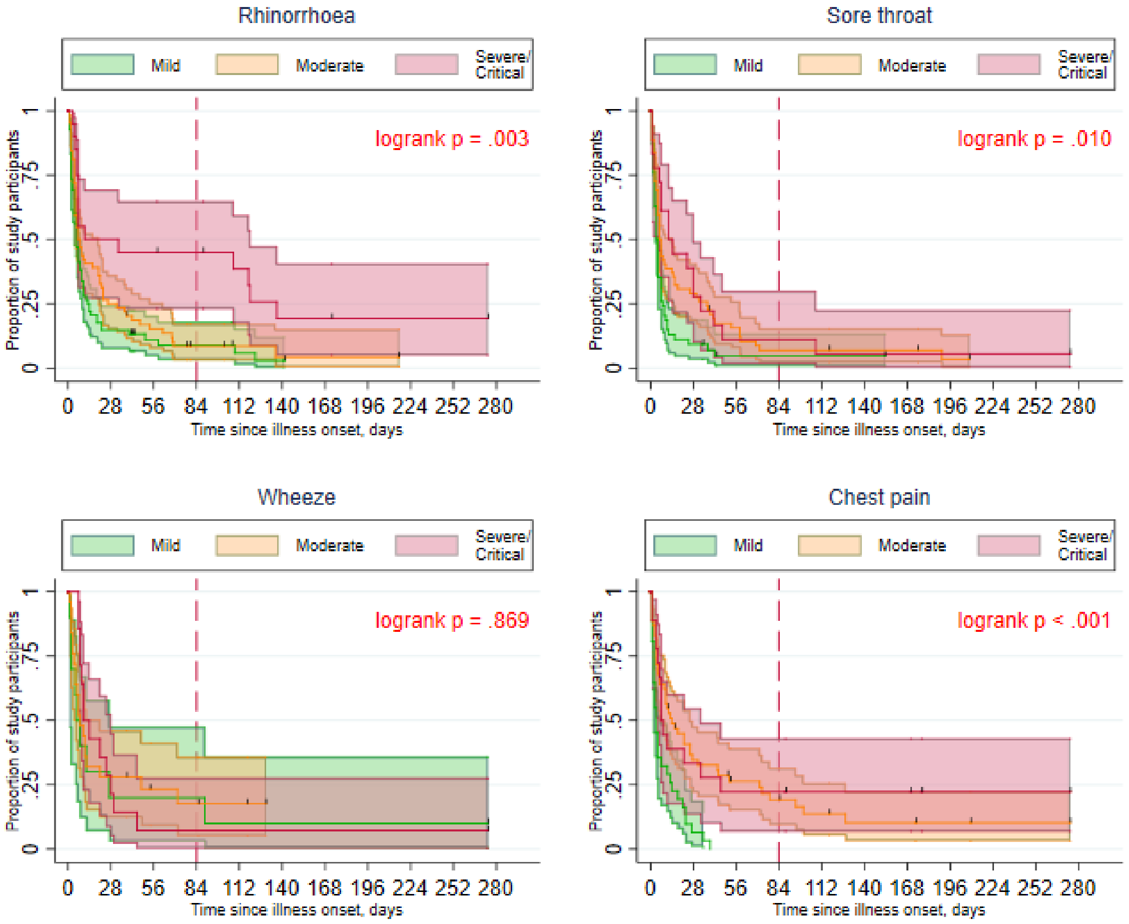

**Figure S3d. Abdominal pain, nausea, diarrhoea and confusion**

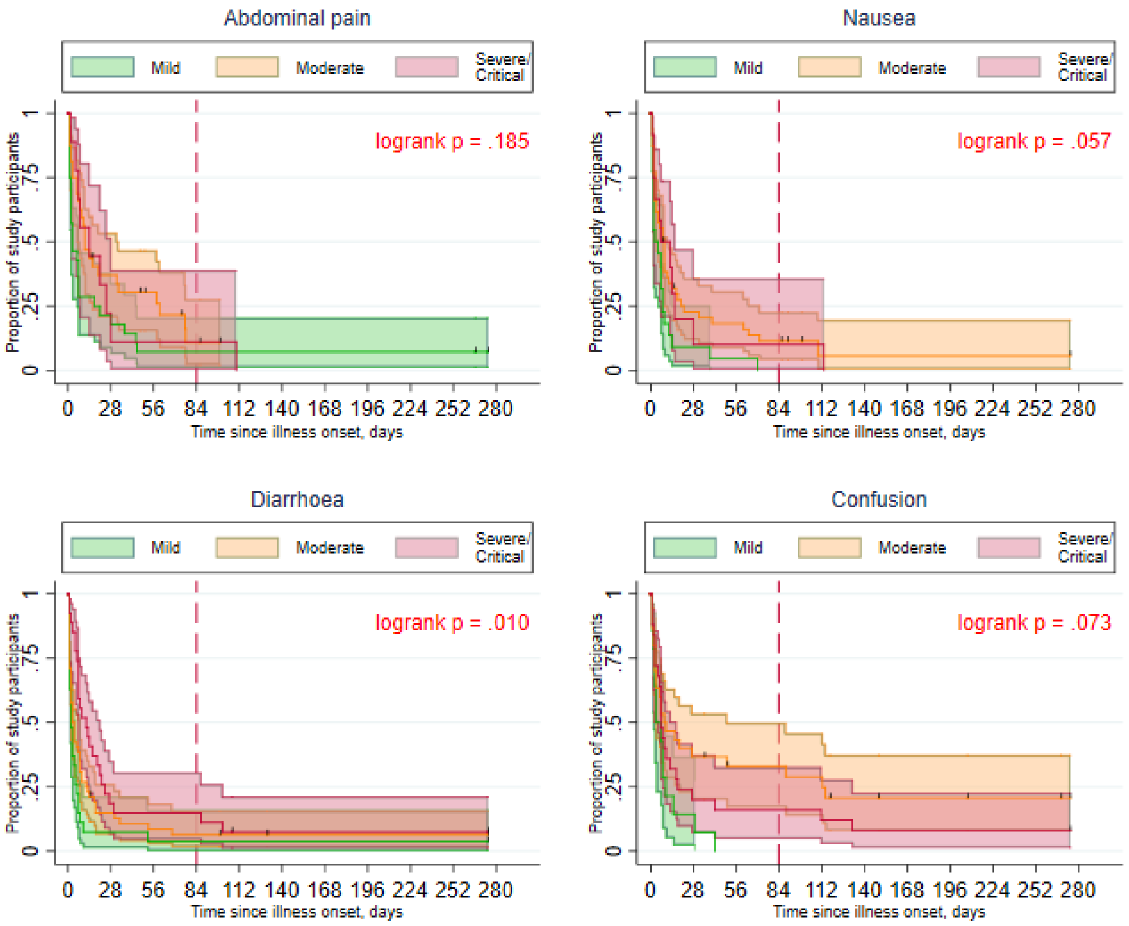

**Figure S3e. Arthralgia and skin rash**

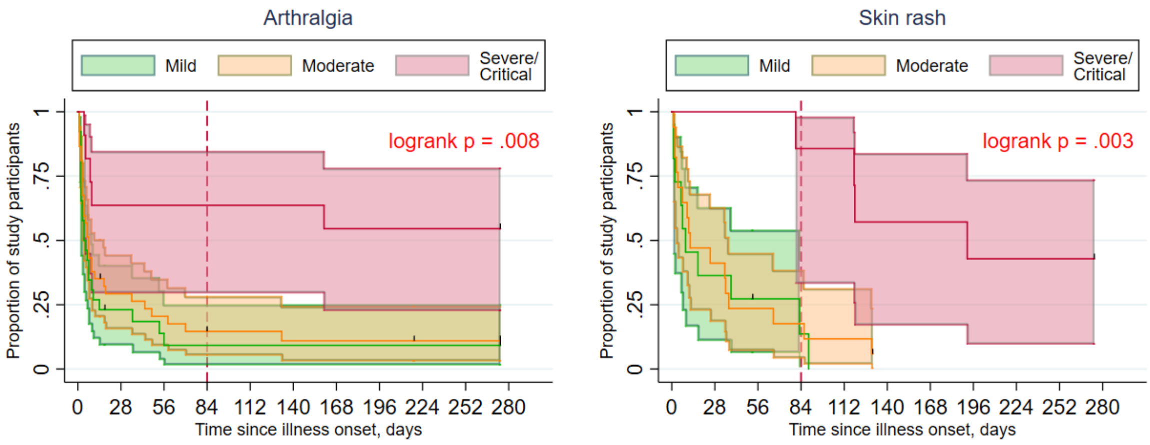

**Figures S4a-b. Sensitivity analyses: Multivariable Cox proportional hazards models of time from illness onset to complete recovery when (a) restricting to prospectively-included participants only and (b) left-truncating data at date of enrolment into the study**

**Figure S4a. Prospectively enrolled participants only**

| Determinants at illness onset | aHR (95%CI) | P-value |
|---|---|---|
| Age, years | | |
| Age (per 10 year increase) | 0.95 (0.81-1.13) | 0.572 |
| Sex | | |
| Male | Ref. | 0.105 |
| Female | 0.72 (0.48-1.07) | |
| BMI, kg/m2 | | |
| Normal weight | Ref. | 0.234 |
| Overweight | 0.76 (0.48-1.22) | |
| Obese | 0.59 (0.29-1.20) | |
| Number of comorbidities | | |
| None | Ref. | 0.543 |
| 1 | 0.84 (0.41-1.73) | |
| 2 | 0.52 (0.20-1.36) | |
| 3 or more | 1.08 (0.23-5.02) | |

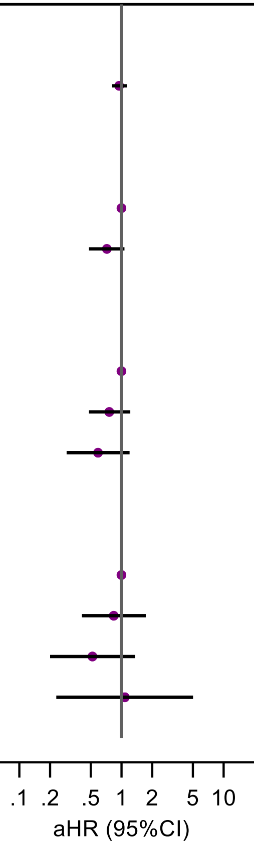


**Figure S4b. Left-truncated at date of enrolment**

| Determinants at illness onset | aHR (95%CI) | P-value |
|---|---|---|
| Age, years | | |
| Age (per 10 year increase) | 1.02 (0.88-1.19) | 0.749 |
| Sex | | |
| Male | Ref. | 0.101 |
| Female | 0.73 (0.49-1.07) | |
| BMI, kg/m2 | | |
| Normal weight | Ref. | 0.200 |
| Overweight | 0.75 (0.48-1.16) | |
| Obese | 0.63 (0.36-1.12) | |
| Number of comorbidities | | |
| None | Ref. | 0.452 |
| 1 | 0.65 (0.34-1.23) | |
| 2 | 0.64 (0.31-1.35) | |
| 3 or more | 0.88 (0.35-2.21) | |

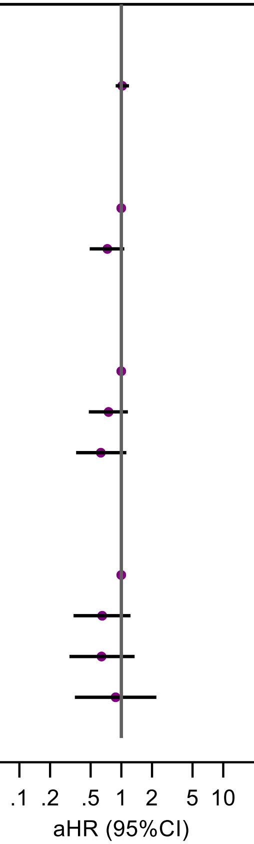


Comorbidities counted are those listed by the WHO as being associated with a higher risk of developing severe or critical COVID-19[11]. Body mass index (BMI) categorised in $kg/m^2$ as: <25, underweight or normal weight; 25-30, overweight; >30, obese. P-value calculated using likelihood ratio test.
